# SARS-CoV-2 Serosurveys: How antigen, isotype and threshold choices affect the outcome

**DOI:** 10.1101/2022.09.09.22279787

**Authors:** Raquel A. Binder, Gavin F. Fujimori, Catherine S. Forconi, George W. Reed, Leandro S. Silva, Priya Saikumar Lakshmi, Amanda Higgins, Lindsey Cincotta, Protiva Dutta, Marie-Claire Salive, Virginia Mangolds, Otuwe Anya, J. Mauricio Calvo Calle, Thomas Nixon, Qiushi Tang, Mireya Wessolossky, Yang Wang, Dominic A. Ritacco, Courtney S. Bly, Stephanie Fischinger, Caroline Atyeo, Peter O. Oluoch, Boaz Odwar, Jeffrey A. Bailey, Ana Maldonado-Contreras, John P. Haran, Aaron G. Schmidt, Lisa Cavacini, Galit Alter, Ann M. Moormann

**Author notes:** Authors contributed equally (Raquel A. Binder, Gavin F. Fujimori, Catherine S. Forconi). Corresponding author: Raquel A. Binder, UMass Chan Medical School, Worcester, MA, 01605, USA.

## Abstract

**Background:** Evaluating the performance of SARS-CoV-2 serological assays and clearly articulating the utility of selected antigen, isotypes and thresholds is crucial to understanding the prevalence of infection within selected communities.

**Methods:** This cross-sectional study, implemented in 2020, screened PCR-confirmed COVID-19 patients (n=86), banked pre-pandemic and negative donors (n=96), health care workers and family members (n=552), and university employees (n=327) for anti-SARS-CoV-2 receptor-binding domain (RBD), trimeric spike protein (S), and nucleocapsid protein (N) IgG and IgA antibodies with a laboratory developed Enzyme-Linked Immunosorbent Assay (ELISA) and tested how antigen, isotype and threshold choices affected the seroprevalence. The following threshold methods were evaluated: (i) mean + 3 standard deviations of the negative controls; (ii) 100% specificity for each antigen/isotype combination; and (iii) the maximal Youden index.

**Results:** We found vastly different seroprevalence estimates depending on selected antigens, isotypes and the applied threshold method, ranging from 0.0% to 85.4%. Subsequently, we maximized specificity and reported a seroprevalence, based on more than one antigen, ranging from 9.3% to 25.9%.

**Conclusions:** This study revealed the importance of evaluating serosurvey tools for antigen, isotype, and threshold-specific sensitivity and specificity, in order to interpret qualitative serosurvey outcomes reliably and consistently across studies.

## Introduction

SARS-CoV-2 seroprevalence studies (i.e. serosurveys) are essential public health tools that can be incorporated into disease spread models to estimate the effective reproductive number, SARS-CoV-2 transmission potentials, disease dynamic forecasts, and assess the impact of public health and clinical interventions, particularly in the beginning of an outbreak [1].

However, given the number of SARS-CoV-2 serosurvey tools, evaluating test performance and clearly articulating their utility across different study populations has become crucial. This is particularly true for laboratory developed serological assays that are not designed for clinical use, were not Emergency Use Authorization (EUA) approved by the FDA, and whose make up and performance vary [2]. Ideally, serosurveys tools are evaluated for their reproducibility, sensitivity and specificity based on PCR-confirmed COVID-19 clinical specimens and well-characterized pre-pandemic negative controls [3]. Further, thresholds for each detected antigen/isotype and potential compound measurements (e.g., final positive call based on two or more SARS-CoV-2 antigens) must be evaluated to allow for qualitative assessments (i.e., providing results that are either positive or negative for the antibodies of interest).

Here, we describe the evaluation and implementation of a laboratory developed SARS-CoV-2 Enzyme-Linked Immunosorbent Assay (ELISA) based on samples from COVID-19 patients, health care workers and their family members, and return to work employees during the first COVID-19 wave in Worcester, Massachusetts, USA from April to August of 2020. Overall, this study aims to demonstrate the importance of (i) rapidly funding academic medical centers to design, evaluate and deploy laboratory developed tests in an outbreak setting, and (ii) evaluating serosurvey tools for antigen, isotype, and threshold-specific sensitivity and specificity, in order to interpret seroprevalence measurements reliably and consistently across study populations.

## Methods

### Study participant recruitment and enrollment

Study participants were enrolled at the University of Massachusetts Chan Medical School (UMass Chan) and the University of Massachusetts Memorial Medical Center (UMass Memorial) in Worcester between April and August of 2020 under the Institutional Review Board (IRB) approved *Consolidated COVID-19 Clinical and Observational Pathogenesis and Epidemiology* (COVID-COPE) Study Protocol (H00020145), see Supplemental Methods for more details. Briefly, once consent was obtained, participants were asked to complete a survey capturing demographic and symptomatic information, and blood was collected. Health care workers (HCW Group) consisted of front-line workers from the UMass Memorial Emergency Department (ED) and their family members (HCW Family Group). Return to work employees (RTW Group), consisted of UMass Chan employees who returned to campus in August after the state mandated stay-at-home order. RTW, HCW and HCW Family members received antibody results (Supplemental Figure 1) with the caveat that this was a research assay and not a diagnostic test. Additionally, patients with COVID-19-like symptoms that were being evaluated at the UMass Memorial ED were approached for study participation (CHC Group). Once consented, the patients or their healthcare proxy completed a survey, and blood was collected. COVID-19 related clinical information, including symptoms, were extracted from the participants’ medical charts. Finally, four sets of banked de-identified banked blood samples (collected between October of 2003 and January of 2020), were sourced from the Moormann Laboratory (n=17) or kindly provided by Dr. Larry Stern (n=13), Dr. Liisa Seline and Dr. Anna Gil from UMass Chan (n=34). Dr. Christopher King from Case Western Reserve University, Cleveland, Ohio (SeroNet U01 CA260539) kindly provided de-identified pre-screened SARS-CoV-2 antibody negative sera samples (Luminex screening for anti-S, N and RB SARS-CoV-2 Wuhan antibodies) collected during a community blood drive in July of 2020 (n=32). Both the pre-pandemic and pre-screened negative samples served as negative controls (total n=96).

### SARS-CoV-2 Enzyme-Linked Immunosorbent Assay (ELISA)

The SARS-CoV-2 ELISA assay developed at the Ragon Institute was implemented at UMass Chan [4], using receptor-binding domain (RBD), S Trimer (S; truncated to 1208 amino acids, deleted transmembrane domain and C-terminal domain), and nucleocapsid protein (N; full-length) SARS-CoV-2 antigens provided by MassBiologics of the UMass Chan, Mattapan, MA [5]. We also used SARS-CoV2 RBD antigen provided by the Ragon Institute, Cambridge, MA. See Supplemental Methods for more details and validation.

### Statistical analysis

Statistical calculations and graphs were done in Prism v8.2.1, R v3.6.1/v4.1.1 and STATA v17. The heatmap was generated by the pheatmap package in R v4.1.1. Three different thresholds were evaluated, see Supplemental Methods for more details. Briefly, we applied the threshold formula “mean + 3 standard deviations of negative controls” *(3SD* threshold) [6]. Next, a maximum specificity threshold was applied by setting the cutoff at 100% specificity for each antigen/isotype combination (i.e., the cutoff value is the highest OD value among the negative control group of each antigen/isotype combination; *Max Spec* threshold). Finally, a threshold based on the maximal Youden index (sensitivity + specificity – 1) on the ROC curve (Supplemental Figure 2) was determined, which balances sensitivity and specificity for each antigen/isotype combination (*Youden* threshold). Experimental samples with OD values equal to or higher than the determined threshold value for each antigen/isotype combination were considered serologically positive (Supplemental Table 1).

## Results

### Study participant recruitment

A total of 212 suspected COVID-19 patients (CHCs) were approached for study participation when they were being evaluated in the ED and 134 (63.2%) were subsequently enrolled. Of those, 86 (64.2%) were PCR-confirmed COVID-19 cases (Table 1). The majority of the CHCs were white (60.4%, n=81) and non-Hispanic (65.7%, n=88), aged 60 years or older (59.7%, n=80; average age 67 years) and male (47.8%, n=64). The latter two determinants being common COVID-19 risk factors and consistent with the demographics of those severely affected during the first wave in Massachusetts [7]. A total of 253 HCWs volunteered to participate in the study, and 299 of their family members were also enrolled. Based on self-reported data, the majority of HCWs were white (76.3%, n=193) and non-Hispanic (80.2%, n=203). The average age of the HCWs was 41 years (range: 21 to 68 years), and 49.8% (n=126) were female. Among the HCW family members who reported demographic information, 54.2% (n=162) were white, 57.5% (n=172) non-Hispanic, and 33.4% (n=100) female. A total of 327 RTW volunteered for the study, of whom the majority were white (70.0%, n=229) and non-Hispanic (93.0%, n=304). About half of the RTWs were female (56.3%, n=184), and the average age was 40 years (range: 22 to 73 years). Hence, a total of 1,013 study participants were successfully enrolled, demographic and symptomatic information recorded, blood samples collected, and sera or plasma screened for anti-SARS-CoV-2 antibodies. Among these study participants, the missing data ranged from 0.3% to 40.8% (Table 1), depending on the demographic category or study population subgroup. Note, this does not include the COVID-19 symptom reports as RTWs, HCWs and HCW family members were only asked to report COVID-19 symptoms if they had a positive SARS-COV-2 PCR test or had reason to believe they were infected. HCW family members were the least likely to answer demographic questions and therefore had the most missing data, despite repeated contact attempts. RTWs were most likely to fill out all the online questionnaires. Part of the draw to the study was the distribution of SARS-CoV-2 antibody reports in the absence of FDA approved tests in the beginning of the pandemic (Supplemental Figure 1).

**Table 1.**
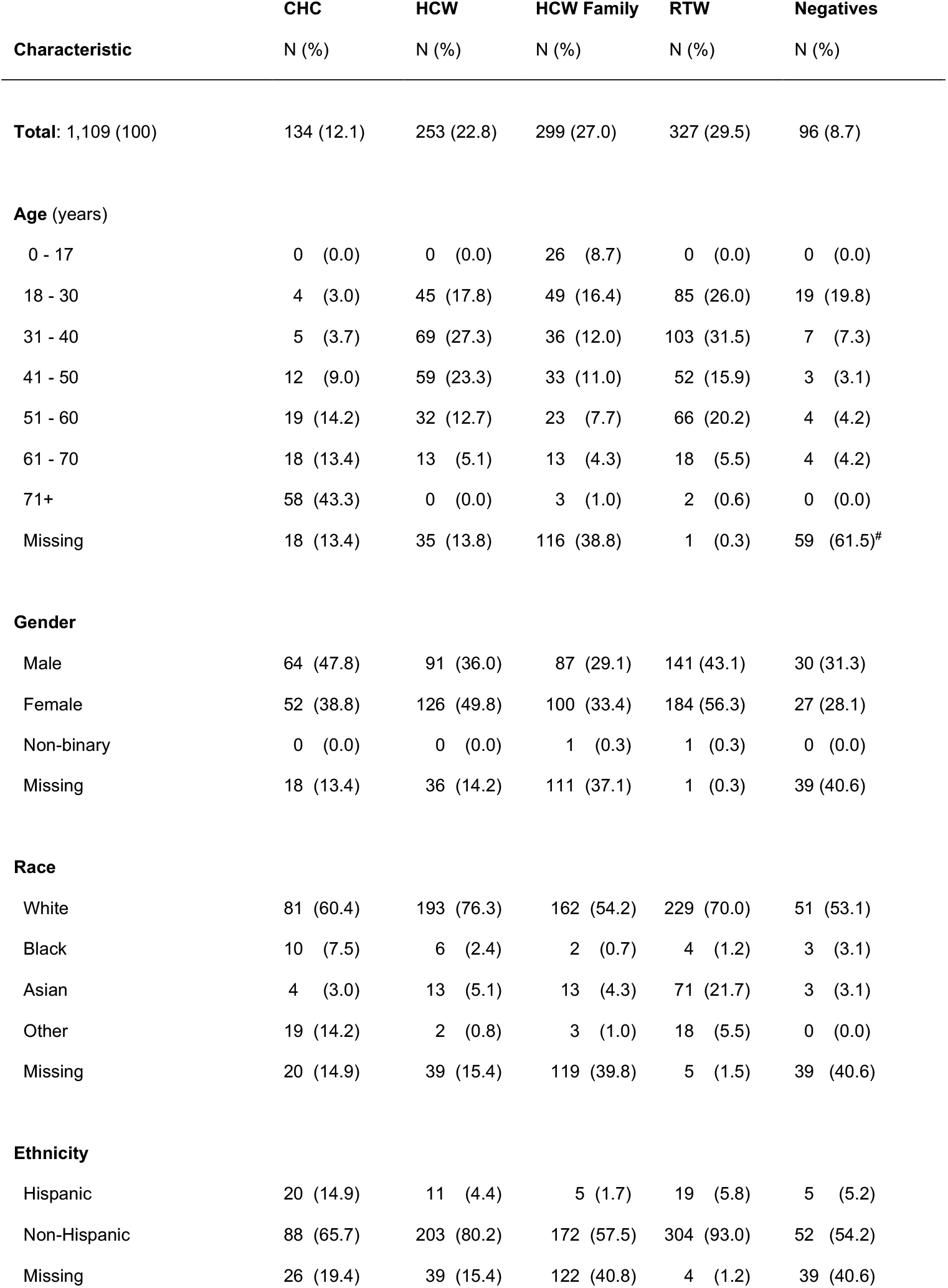
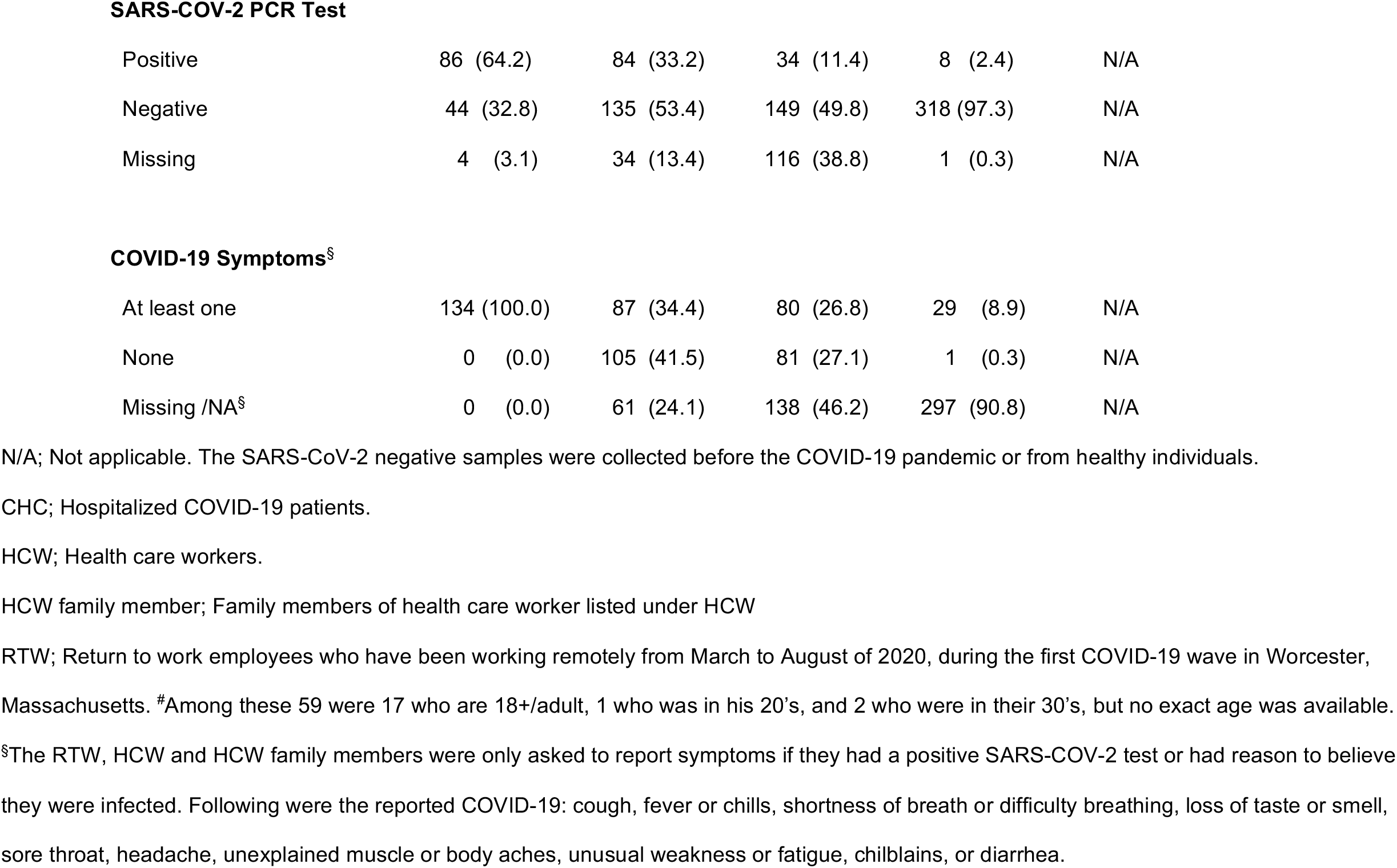
Demographics, SARS-CoV-2 PCR results and symptom distribution among study participants (n=1,109).

| Characteristic | CHC<br>N (%) | HCW<br>N (%) | HCW Family<br>N (%) | RTW<br>N (%) | Negatives<br>N (%) |
| --- | --- | --- | --- | --- | --- |
| **Total: 1,109 (100)** | 134 (12.1) | 253 (22.8) | 299 (27.0) | 327 (29.5) | 96 (8.7) |
| **Age (years)** |  |  |  |  |  |
| 0 - 17 | 0 (0.0) | 0 (0.0) | 26 (8.7) | 0 (0.0) | 0 (0.0) |
| 18 - 30 | 4 (3.0) | 45 (17.8) | 49 (16.4) | 85 (26.0) | 19 (19.8) |
| 31 - 40 | 5 (3.7) | 69 (27.3) | 36 (12.0) | 103 (31.5) | 7 (7.3) |
| 41 - 50 | 12 (9.0) | 59 (23.3) | 33 (11.0) | 52 (15.9) | 3 (3.1) |
| 51 - 60 | 19 (14.2) | 32 (12.7) | 23 (7.7) | 66 (20.2) | 4 (4.2) |
| 61 - 70 | 18 (13.4) | 13 (5.1) | 13 (4.3) | 18 (5.5) | 4 (4.2) |
| 71+ | 58 (43.3) | 0 (0.0) | 3 (1.0) | 2 (0.6) | 0 (0.0) |
| Missing | 18 (13.4) | 35 (13.8) | 116 (38.8) | 1 (0.3) | 59 (61.5)<sup>#</sup> |
| **Gender** |  |  |  |  |  |
| Male | 64 (47.8) | 91 (36.0) | 87 (29.1) | 141 (43.1) | 30 (31.3) |
| Female | 52 (38.8) | 126 (49.8) | 100 (33.4) | 184 (56.3) | 27 (28.1) |
| Non-binary | 0 (0.0) | 0 (0.0) | 1 (0.3) | 1 (0.3) | 0 (0.0) |
| Missing | 18 (13.4) | 36 (14.2) | 111 (37.1) | 1 (0.3) | 39 (40.6) |
| **Race** |  |  |  |  |  |
| White | 81 (60.4) | 193 (76.3) | 162 (54.2) | 229 (70.0) | 51 (53.1) |
| Black | 10 (7.5) | 6 (2.4) | 2 (0.7) | 4 (1.2) | 3 (3.1) |
| Asian | 4 (3.0) | 13 (5.1) | 13 (4.3) | 71 (21.7) | 3 (3.1) |
| Other | 19 (14.2) | 2 (0.8) | 3 (1.0) | 18 (5.5) | 0 (0.0) |
| Missing | 20 (14.9) | 39 (15.4) | 119 (39.8) | 5 (1.5) | 39 (40.6) |
| **Ethnicity** |  |  |  |  |  |
| Hispanic | 20 (14.9) | 11 (4.4) | 5 (1.7) | 19 (5.8) | 5 (5.2) |
| Non-Hispanic | 88 (65.7) | 203 (80.2) | 172 (57.5) | 304 (93.0) | 52 (54.2) |
| Missing | 26 (19.4) | 39 (15.4) | 122 (40.8) | 4 (1.2) | 39 (40.6) |
| **SARS-COV-2 PCR Test** |  |  |  |  |  |
| Positive | 86 (64.2) | 84 (33.2) | 34 (11.4) | 8 (2.4) | N/A |
| Negative | 44 (32.8) | 135 (53.4) | 149 (49.8) | 318 (97.3) | N/A |
| Missing | 4 (3.1) | 34 (13.4) | 116 (38.8) | 1 (0.3) | N/A |
| **COVID-19 Symptoms<sup>§</sup>** |  |  |  |  |  |
| At least one | 134 (100.0) | 87 (34.4) | 80 (26.8) | 29 (8.9) | N/A |
| None | 0 (0.0) | 105 (41.5) | 81 (27.1) | 1 (0.3) | N/A |
| Missing /NA<sup>§</sup> | 0 (0.0) | 61 (24.1) | 138 (46.2) | 297 (90.8) | N/A |

N/A; Not applicable. The SARS-CoV-2 negative samples were collected before the COVID-19 pandemic or from healthy individuals.
CHC; Hospitalized COVID-19 patients.
HCW; Health care workers.
HCW family member; Family members of health care worker listed under HCW
RTW; Return to work employees who have been working remotely from March to August of 2020, during the first COVID-19 wave in Worcester, Massachusetts. <sup>#</sup>Among these 59 were 17 who are 18+/adult, 1 who was in his 20's, and 2 who were in their 30's, but no exact age was available.
<sup>§</sup>The RTW, HCW and HCW family members were only asked to report symptoms if they had a positive SARS-COV-2 test or had reason to believe they were infected. Following were the reported COVID-19: cough, fever or chills, shortness of breath or difficulty breathing, loss of taste or smell, sore throat, headache, unexplained muscle or body aches, unusual weakness or fatigue, chilblains, or diarrhea.

### Threshold methods and sensitivity/specificity

Three different threshold methods were evaluated to convert the quantitative OD values into qualitative positive/negative results and subsequent sensitivity and specificity: the 3 SD, Max Spec and Youden threshold. PCR-confirmed SARS-CoV-2 positive CHCs (n=86) and negative samples (n=96) were screened for anti-RBD, anti-S and anti-N SARS-CoV-2 IgA and IgG antibodies by ELISA. We further evaluated (i) combining the three antigen/isotype combinations (RBD IgG, N IgG, and RBD IgA) with the largest area under the ROC curve (i.e., with maximum sensitivity and specificity; Top Three); (ii) combining all IgA isotypes (N, RBD and S IgA; All IgA); and (iii) combining all IgG isotypes (N, RBD and S IgG; All IgG) as serological outcome measures (Figure 1). We used isotype-based compound measurements since a heatmap of CHC, HCW and RTW OD values with at least one positive antigen/isotype combination clustered by isotype, rather than antigen (Figure 2). The clustering was confirmed with a correlogram comparing antigen/isotype combinations. Again, when analyzing PCR-confirmed CHCs or all seropositive participants, the strongest positive correlation was between the same isotypes rather than the same antigens (Supplemental Figure 3). This clustering of IgG and IgA responses to SARS-CoV-2 infection has been described by others [8, 9], and is likely a function of time since infection [10] and protective class-switching [8].

**Figure 1.**
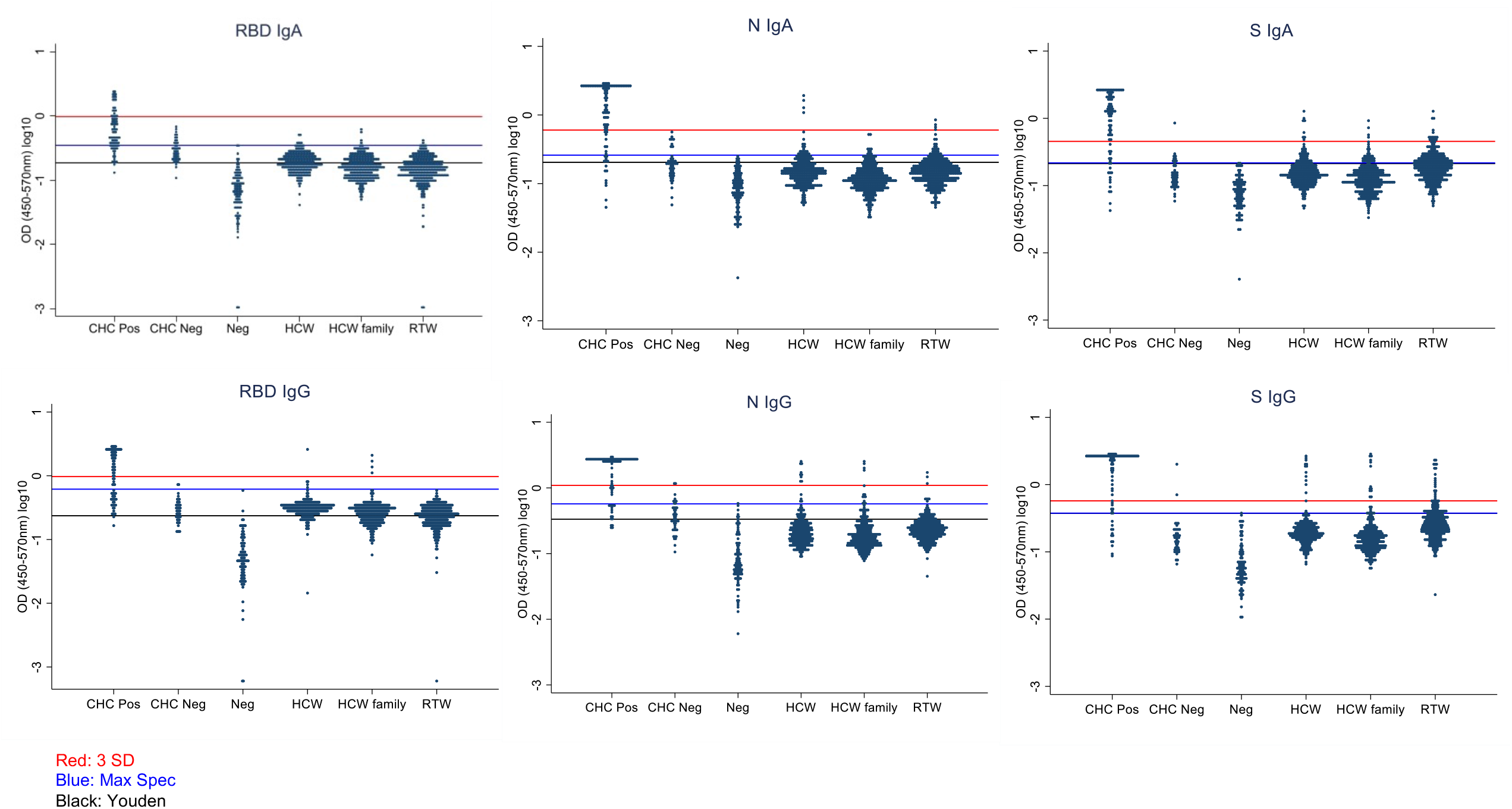
Optical density value distribution among each measured subgroup. Optical density values (450-570nm) were plotted according to each antigen/isotype combination and subgroup (y-axis in log10 scale). The red line indicates the cut off for the 3 standard deviation above the mean threshold method (3 SD). The blue line indicates the cut off when the threshold was chosen at the highest value of the negative controls (Max Spec). The black line indicates the cut off when the threshold was chosen based on the Youden threshold (Youden). RBD; SARS-CoV-2 Receptor-Binding Domain. S Trimer; SARS-CoV-2 Spike Trimer. N; SARS-CoV-2 Nucleocapsid Protein. CHC; Hospitalized COVID-19 patients. HCW; Health care workers. HCW family member; Family members of health care worker listed under HCW. RTW; Return to work employees who had been working remotely from March to August of 2020, during the first COVID-19 wave in Worcester, Massachusetts.

**Figure 2.**
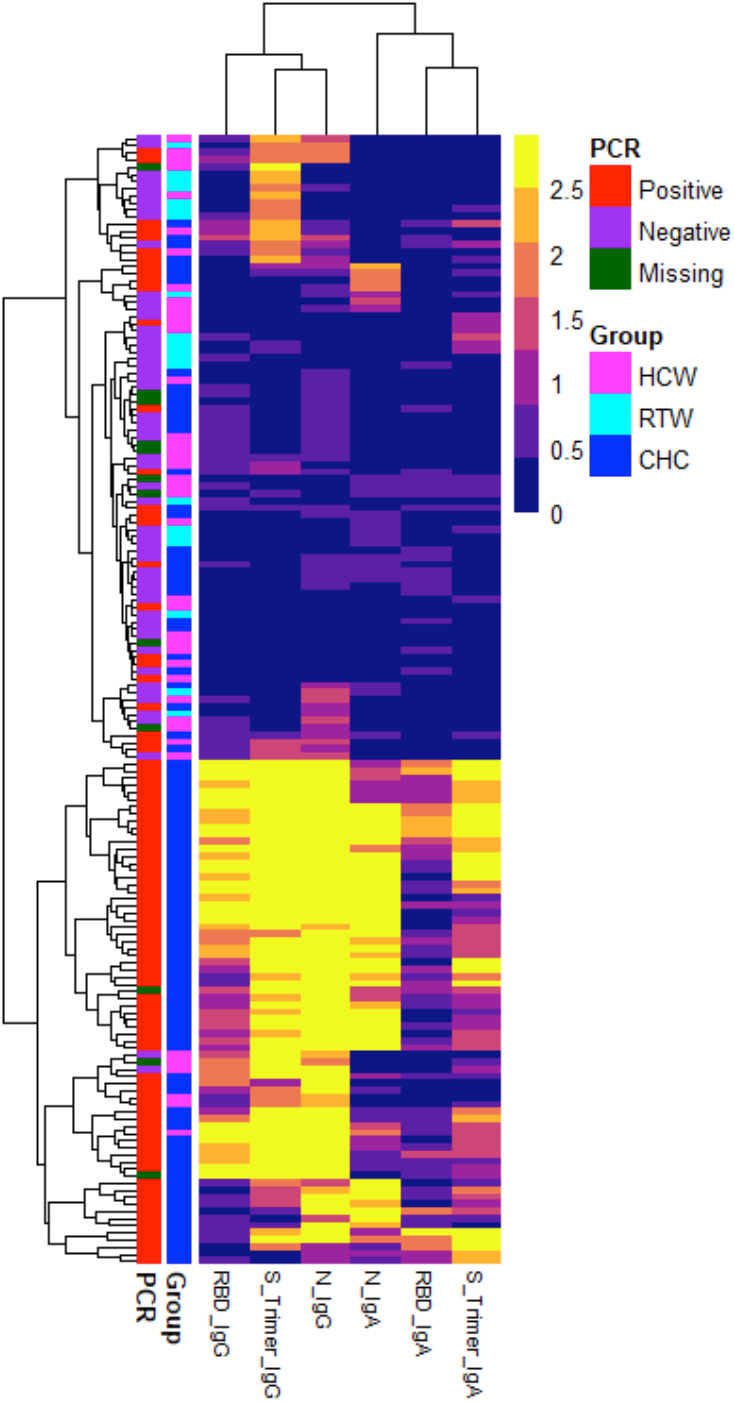
Heatmap of optical density values. A heatmap was built based on unbiased clustering of the optical density (OD) values of the experimental subgroups with at least one positive antigen/isotype combination. The results clustered by isotype, rather than antigen (see top branching into IgG and IgA from left to the right). RBD; SARS-CoV-2 Receptor-Binding Domain. S Trimer; SARS-CoV-2 Spike Trimer. N; SARS-CoV-2 Nucleocapsid Protein. CHC; Hospitalized COVID-19 patients. HCW; Health care workers and their family members. RTW; Return to work employees who had been working remotely from March to August of 2020, during the first COVID-19 wave in Worcester, Massachusetts.

Applying the 3 SD threshold resulted in 100% specificity for all isotype/antigen combinations across IgG and IgA (Table 2). Conversely, sensitivity was low and ranged from 25.6% to 76.7%. Sensitivities for RBD IgA and IgG, 25.6% and 55.8%, were particularly low. Sensitivities for IgA and IgG S Trimer and IgA and IgG N were similar and ranged from 72.1% to 76.7%. The RBD IgM antigen/isotype combination resulted in relatively low sensitivity (67.4%) and specificity (98.5%), similar to previously published results [4, 11]. Given the quick resolution of IgM, while appearing concomitantly with IgG after SARS-CoV-2 infection [3], IgM was not deemed a useful serosurvey tool and was omitted from the rest of the analyses.

**Table 2.**
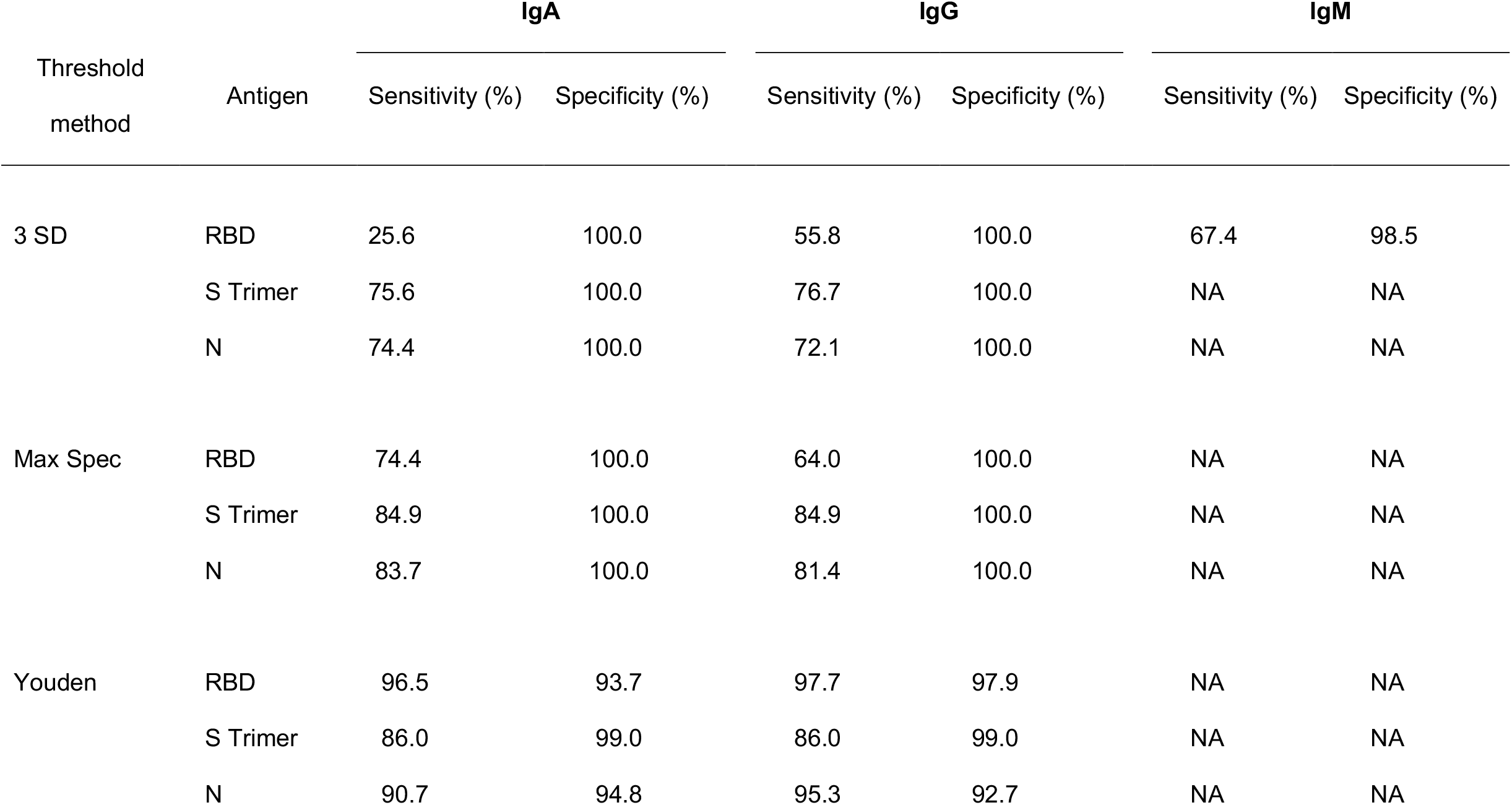
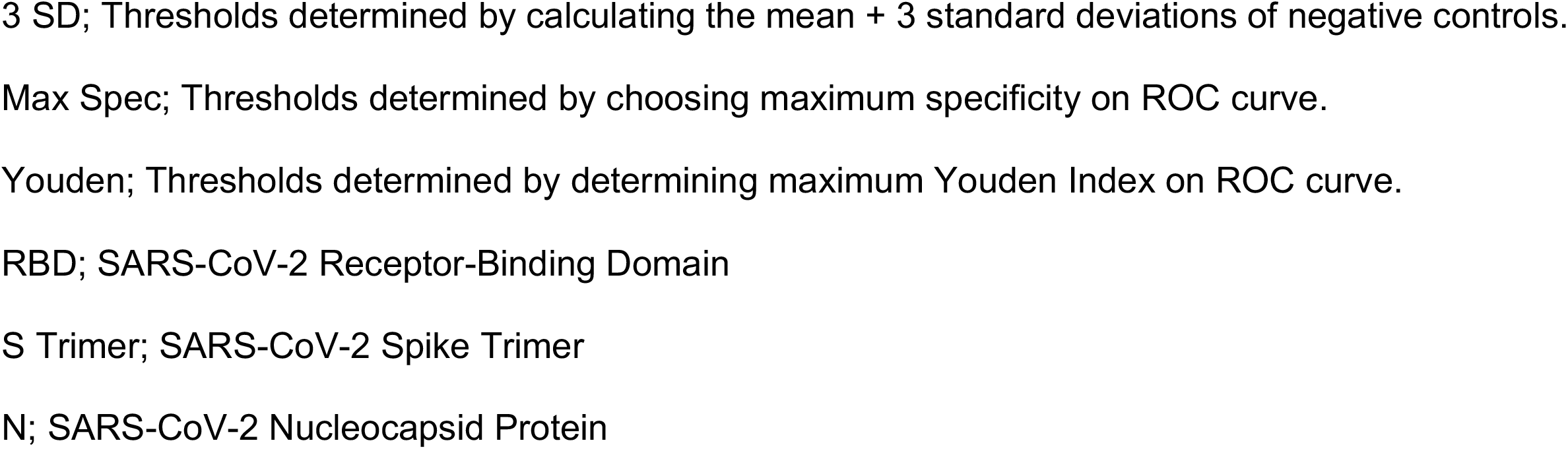
Sensitivity and specificity for each threshold method and isotype/antigen combination based on PCR-confirmed and hospitalized COVID-19 patients (n=86) and negative samples (n=96).

Based on the Max Spec threshold (fixing the specificity at 100%), sensitivity for RBD IgA and IgG increased compared to the 3 SD threshold but remained relatively low, 74.4% and 64.0%, respectively. Again, sensitivities for IgA and IgG S Trimer and IgA and IgG N were similar and ranged from 81.4% to 84.9%

Based on the Youden threshold, robust sensitivity was obtained across all antigen/isotype combinations, ranging from 86.0% to 97.7%. Sensitivity for RBD IgA and IgG were 96.5% and 97.7%, respectively. Sensitivities for IgA and IgG S Trimer and IgA and IgG N ranged from 86.0% to 95.3%. Conversely, specificity was slightly lower compared to the other two methods and ranged from 92.7% to 99.0%, see Table 2.

### HCW, HCW Family and RTW serological results

Next, we screened HCW (n=253), HCW Family members (n=299), and RTWs (n=327) for anti-RBD, anti-S and anti-N SARS-CoV-2 IgA and IgG antibodies by ELISA. The overall OD value spread for the HCW, HCW Family and RTW subgroups were similar (Figure 1). However, the overlap between the PCR-positive CHCs (positive controls) and negative control values varied notably depending on the antigen/isotype combination, affecting the cutoffs for each method and therefore the subsequently determined seroprevalence for each subgroup. Furthermore, most negative control OD values were notably lower than the experimental subgroups (HCW, HCW Family, and RTW) for RBD IgG, S IgG, and N IgG, but not so much for the IgA equivalents. Hence, when applying the three different thresholds to the experimental subgroups the seroprevalence outcomes varied widely, not just across threshold methods (Youden, 3 SD, and Max Spec), but also across antigen/isotype combinations (Figure 1, Figure 3 and Supplemental Figure 4).

**Figure 3.**
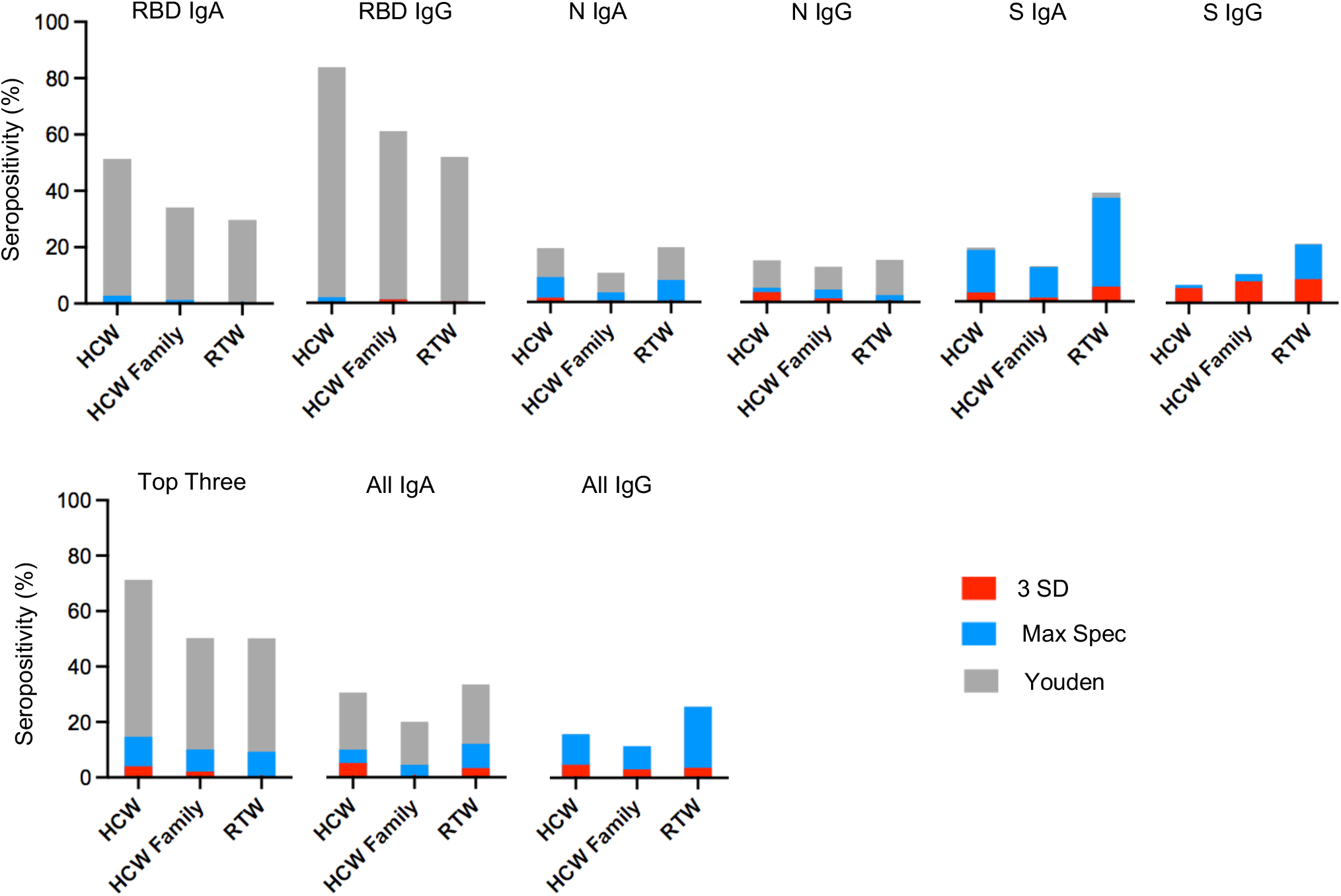
Seropositivity of each subgroup, according to the antigen/isotype combination. The red bars graph the seroprevalence based on the cut off for the 3 standard deviation above the mean threshold method (3 SD). The blue bars graph the seroprevalence based on the maximum specificity threshold (Max Spec). The black bars graph the seroprevalence based on the Youden threshold (Youden). RBD; SARS-CoV-2 Receptor-Binding Domain. S Trimer; SARS-CoV-2 Spike Trimer. N; SARS-CoV-2 Nucleocapsid Protein. HCW; Health care workers. HCW family member; Family members of health care worker listed under HCW. RTW; Return to work employees who had been working remotely from March to August of 2020, during the first COVID-19 wave in Worcester, Massachusetts.

When comparing the overall seroprevalence for each antigen/isotype combination, RBD IgA, RBD IgG and the Top Three compound estimate (RBD IgG, RBD IgA, and N IgG) yielded the highest seroprevalence, especially when applying the Youden threshold, ranging from 29.7% to 85.4% (Supplemental Table 2, Figure 3). The S IgA- and All IgA-based seroprevalence ranged from 12.5% to 38.6% based on the Youden threshold (Supplemental Table 2). Overall, N IgA-, N IgG- and S IgG-based seroprevalence was lowest, ranging from 0.0% to 20.6%, independent of applied threshold; again, noting the wide range of determined seroprevalence (Supplemental Table 2).

When comparing the three participant sub-groups based on the Youden threshold, the HCWs had the highest seroprevalence followed by the HCW Family and RTWs groups when measuring RBD IgA (51.4%, 34.1%, and 29.7%, respectively) and RBD IgG (85.4%, 62.2%, and 52.9%, respectively; Supplemental Table 2 and Figure 3). For the other antigen/isotype combinations, the order was less pronounced, except for S IgA where the RTW subgroup had the highest seroprevalence based on the Youden and Max Spec thresholds, compared to the HCWs and HCW Family subgroups.

When comparing the threshold methods, applying the Youden cutoff, which was the most sensitive one, yielded the highest seroprevalence across all antigen/isotype combinations even though with a wide range (6.0% to 85.4%; Supplemental Table 2, Figure 3). Further, while the RBD IgA-, RBD IgG- and the Top Three compound-based seroprevalence estimates were highest when applying the Youden threshold method, they also represented the most notable discrepancy when comparing the Youden to the Max Spec and 3 SD thresholds. Based on the Youden threshold method, the RBD IgG-based seroprevalence among all three subgroups ranged from 52.9% to 85.4%, as compared to the 3 SD and Max Spec thresholds where the seroprevalence ranged from 0.0% to 2.0%.

Given the notable variation in OD spread among the antigen/isotype combinations and the resulting wide range of seroprevalence estimates (including the overinflated seroprevalence based on RBD IgA/IgG when applying the Youden threshold, Figure 3), maximizing the specificity and including more than one antigen (compound measurements) for the overall serosurvey measurement was prioritized.

Being compound measurements with the least antigen/isotype-dependent variation, the Top Three and the All IgG compound estimates, based on the Max Spec threshold, were considered the most reliable outcome measurements to determine seroprevalence among our study population. Based on the Top Three compound estimate and the Max Spec threshold, we estimated that the seroprevalence among the HCW, HCW Family, and RTW study participants was 14.7%, 10.1%, and 9.3%, respectively (HCWs ranking highest; Supplemental Table 2, Figure 3). Based on the All IgG compound estimate and the Max Spec threshold, we estimated that the seroprevalence among the HCW, HCW Family, and RTW study participants was 15.9%, 11.5%, and 25.9%, respectively (RTWs ranking highest). Hence, based on these two compound estimates and the Max Spec threshold, we estimate that the anti-SARS-CoV-2 antibody seroprevalence among the HCW, HCW Family, and RTW study participants ranged from 9.3% to 25.9% during the first COVID-19 wave in the Spring of 2020, while the ranking was unclear due to relatively low overall seroprevalence. Note that at the time, no COVID-19 vaccines were available and therefore detecting any SARS-CoV-2 antibodies was considered an indication of infection.

Finally, participant COVID-19 symptoms and test reports were analyzed in order to validate the serological results. As part of the participant surveys, HCWs and HCW Family members, and RTWs were asked to report COVID-19-like symptoms if they either had a positive SARS-CoV-2 PCR test or if they thought they had been infected (Table 1). Among HCWs, HCW family members and RTW study participants, HCWs reported the most positive SARS-CoV-2 PCR tests (33.2%, n=84) and COVID-19 symptoms (34.4%, n=87), followed by HCW family members and RTWs (Table 1). Thus, confirming a potential HCW, HCW Family and RTW ranking. Interestingly, among the RTWs, HCWs, and HCW family members who reported SARS-CoV-2 PCR positive tests only 54.8% (n=69) reported COVID-19 symptoms (32.5%, n=41 reported no symptoms and 12.7%, n=16 did not answer that question). Note, molecular SARS-CoV-2 tests were not widely available outside of hospital settings during the study period.

## Discussion

Given the number of serological SARS-CoV-2 assays, evaluating test performance and ideal cutoffs, along with clearly articulating their utility across different study populations is crucial. Here, we validated a laboratory developed SARS-CoV-2 ELISA and screened ED patients with COVID-19 symptoms (CHC Group), HCWs (HCW Group), their family members (HCW Family Group), and return to work employees (RTW Group) for anti-RBD, -S, and -N IgG and IgA antibodies. Further, we evaluated three different methods to transform quantitative OD values into qualitative results and seroprevalence estimates.

The Youden index is a summary measure of the ROC curve designed to provide optimal separation of negative and positive values and therefore maximize sensitivity and specificity. The Max Spec and the 3 SD are thresholds designed to maximize specificity and are based on negative/pre-pandemic samples only. The present study found notable variation in seroprevalence estimates comparing the three threshold methods, ranging from 0.0% to 85.4%. The seroprevalence outcomes were the lowest and varied least across antigen/isotype combination based on the 3 SD threshold. While applying the Youden threshold increases sensitivity, important in a clinical setting, it yielded unrealistically high seroprevalence estimates of up to 85.4% (RBD IgG, HCW) that varied significantly among antigen/isotype combinations in our case. For example, the RBD IgG-based seroprevalence among the HCW, HCW Family an RTW groups ranged from 52.9% to 85.4% when applying the Youden threshold and from 0.0% to 2.0% when applying the Max Spec threshold (Supplemental Table 2). While such discrepancies were most drastic among RBD-based seroprevalence estimates and were unrealistically high for both RBD IgA and IgG when applying the Youden method, the seroprevalence outcomes were not as different when comparing the three thresholds for N IgA, N IgG, and S IgG. This could have been due to variation in antigen/isotype specific immune responses among the PCR-confirmed CHC, prep-pandemic and negative participants, or the respective half-life of circulating antibodies. For example, *Bolotin et al*. found a substantial decline in anti-N SARS-CoV-2 antibodies over a 5-month period as part of a Canadian serosurveillance program [12]. Further, others have shown that S-specific serum IgA levels decay significantly (p < 0.002) faster than S-specific IgG post-COVID-19 mRNA vaccination [10]. Hence, the choice of (i) measured antigen/isotype, (2) threshold method, (3) presumably positive and negative controls will affect the threshold and therefore the qualitative outcomes of a serological assays notably.

These results emphasized the importance of maximizing assay specificity, as generally done in population-based studies, and using compound measurements for our final serosurvey outcome. For this study, the Top Three and the All IgG compound estimates were considered the most reliable, along with the Max Spec threshold. Based on these two compound estimates and the Max Spec threshold, we estimated that the anti-SARS-CoV-2 antibody seroprevalence among the HCW, HCW Family, and RTW study participants ranged from 9.3% to 25.9% during the first COVID-19 wave in the Spring of 2020, while the ranking was unclear due to relatively low overall seroprevalence. A similar hospital-based study found seroprevalence estimates of 5.5% among HCWs in Boston, Massachusetts in July of 2020 [13]. Another Massachusetts population-based study found seroprevalence estimates of 31.5% in April of 2020 [14].

However, the difference could be due to the serosurvey tool as the study used a IgM-IgG point-of-care lateral flow immunoassay. A US-wide blood donation-based SARS-CoV-2 seroprevalence study found that infection-induced seroprevalence estimates increased from 3.5% to 11.5% between July and December of 2020, and estimated the northeast (including MA) to have reached a seroprevalence of 19.3% by May 2021 when the study ended [15]. Hence, our HCW, HCW Family, and RTW populations, fall into the estimated ranges in similar time periods.

Returning antibody results (as means to determine the likelihood of a past infection) turned out to be a welcomed benefit to study participation in the absence of widely available clinically certified diagnostic tests at the time. We made it clear that the antibody reports were not based on a certified diagnostic assay but rather on a laboratory developed test and should not be interpreted as demonstrating protection from subsequent SARS-CoV-2 infections.

The study had limitations. For example, our study participants were mostly white and non-Hispanic. Further, our results may not be applied to currently circulating SARS-CoV-2 variants since the study samples were collected during the early phase of the pandemic.

Overall, there was a general lack of molecular tests during active infection for non-hospitalized cases. Hence, we were not able to confirm whether our study included asymptomatic infections or potential non-seroconverters and whether the lack of antibodies may have been a function of time since infection. Similarly, there was only limited information on days since symptom onset for the CHCs which would have allowed determining sensitivity and specificity in terms of time since symptom onset. Further, there could have been self-selection bias among HCW, HCW Family and RTW whereby people who thought they had been infected or exposed were more likely to enroll in our study and therefore artificially inflate the seroprevalence. Finally, ELISAs are being increasingly replaced by multiplex bead assays. However, methods to define seropositivity still need to be applied [16]. Note, in the COVID-19 vaccine era, serological assays are designed to distinguish between vaccination (i.e., anti-S or -RBD only antibodies) and infection (i.e. anti-S, -RBD and/or -N antibodies), even though one has to consider potential cross-reactivity with endemic human CoVs, especially OC43 and HKU1 which are most closely related to SARS-CoV-2 [17].

In summary, the present study found notable variation among seroprevalence outcomes depending on the antigen/isotype combination and the chosen threshold method. Robust serosurveys are powerful public health tools that can determine the extent of previous SARS-CoV-2 infections or vaccination rates among populations with different exposure risks. Knowing the fraction of susceptible individuals among a specific population assists with (i) establishing effective risk assessments, (ii) implementing specific infection control measures, and (iii) determining the effectiveness of those measures over time. This is especially important in regions outside the US, where vaccinations are not widely implemented and longitudinal serosurveys will help monitor changes in transmission patterns. However, to obtain reliable seroprevalence estimates, serosurvey tools need to be evaluated for antigen, isotype, and threshold-specific sensitivity and specificity, in order to interpret qualitative serosurvey outcomes reliably and consistently across study populations.

## Data Availability

All data produced in the present study are available upon reasonable request to the authors

## Acknowledgement

We thank the study participants and their families for giving us their time and attention during extraordinary times, without their participation the study would not have been possible. We thank Dr. Larry Stern, Dr. Liisa Seline, and Dr. Anna Gil from UMass Chan and Dr. Christopher King from Case Western Reserve University for providing pre-pandemic and negative serological control samples. We thank Tasso Inc. for the donation of kits at the onset of this study.

